# Investigating the impact of physical activity on mitochondrial function in Parkinson’s disease (PARKEX): Study protocol for a randomised controlled clinical trial

**DOI:** 10.1101/2023.10.20.23297305

**Authors:** Juan Carlos Magaña, Cláudia M. Deus, Laura Baldellou, M Avellanet, E Gea-Rodríguez, Silvia Enriquez Calzada, Ariadna Laguna, Marta Martínez-Vicente, Jorge Hernández-Vara, Maria Giné-Garriga, Susana P. Pereira, Joel Montané

**Affiliations:** Facultat de Psicologia, Ciències de l’Educació i de l’Esport Blanquerna, Ramon Llull University, Barcelona, Spain; CNC-UC, Center for Neuroscience and Cell Biology, University of Coimbra, Coimbra, Portugal; CIBB, Center for Innovative Biomedicine and Biotechnology, University of Coimbra, Coimbra, Portugal; Facultat de Ciències de la Salut, Blanquerna, Ramon Llull University, Barcelona, Spain; Hospital Nostra Senyora de Meritxell, Escaldes-Engordany, Andorra; Universitat d’Andorra, Sant Julià de Lòria, Andorra; Grup de Malalties Neurodegeneratives de la Vall d’Hebron. Vall d’Hebron Institut de Recerca (VHIR), Barcelona, Spain; Aligning Science Across Parkinson’s (ASAP) Collaborative Research Network; Chevy Chase, MD 20815, USA; Institut de Neurociències-Autonomous University of Barcelona (INc-UAB); 08193 Cerdanyola del Vallès, Spain; Laboratory of Metabolism and Exercise (LaMetEx), Research Centre in Physical Activity, Health and Leisure (CIAFEL), Laboratory for Integrative and Translational Research in Population Health (ITR), Faculty of Sports, University of Porto, Porto, Portugal

**Author notes:** these authors contributed equally to this work (Juan Carlos Magaña, Cláudia M. Deus). these authors contributed equally to this work (Susana P. Pereira, Joel Montané). Correspondence should be addressed to: Dr. Joel Montané, Blanquerna School of Health Science, Universitat Ramon Llull Carrer de Padilla, 326-332, 08025 Barcelona, Spain, Dr. Susana P. Pereira, CNC-Center for Neuroscience and Cell Biology, University of Coimbra, Coimbra, Portugal.

**Keywords:** Physical activity, Exercise therapy, Biomarker, Neuroprotection, Mitochondrial diseases, Parkinson’s disease

## Abstract

Parkinson’s disease (PD) is characterized by the progressive dopaminergic neuron degeneration, resulting in striatal dopamine deficiency. Mitochondrial dysfunction and oxidative stress are associated with PD pathogenesis. Physical activity (PA) has been shown to ameliorate neurological impairments and to impede age-related neuronal loss. In addition, skin fibroblasts have been identified as surrogate indicators of pathogenic processes correlating with clinical measures. The PARKEX study aims to compare the effects of two different PA programs, analyzing the impact on mitochondrial function in patients’ skin fibroblasts as biomarkers for disease status and metabolic improvement. Early-stage PD patients (n=24, H&Y stage I to III) will be randomized into three age– and sex-matched groups. Group 1 (n=8) will undergo basic physical training (BPT) emphasizing strength and resistance. Group 2 (n=8) will undergo BPT combined with functional exercises (BPTFE), targeting the sensorimotor pathways that are most affected in PD (proprioception-balance-coordination) together with cognitive and motor training (Dual task training). Group 3 (n=8) will serve as control (sedentary group; Sed). Participants will perform three sessions per week for 12 weeks. Assessment of motor function, quality of life, sleep quality, cognitive aspects and humor will be conducted pre– and post-intervention. Patient skin fibroblasts will be collected before and after the intervention and characterized in terms of metabolic remodeling and mitochondrial bioenergetics. Ethical approval has been given to commence this study. This trial is registered at clinicaltrials.gov (NCT05963425)

## Introduction

Parkinson’s disease (PD) is a disorder characterized by the progressive degeneration of dopaminergic neurons resulting in a deficiency of dopamine in the *striatum* in the basal ganglia [1]. PD patients experience both motor and non-motor symptoms, including anxiety, depression, sleep and gastrointestinal alterations, among others. The motor symptoms typically manifest gradually, initially asymmetrical and later bilateral. The most common primary motor symptoms include *akinesia* (lack of spontaneous voluntary movement), *bradykinesia* (slowness of movement), resting tremor (hands, arms, legs, jaw, and face), rigidity (arms, legs, and trunk), and postural instability. Additionally, people with PD may experience difficulties with balance and coordination [2]. Although the exact mechanism behind the neurodegeneration of dopaminergic cells remains uncertain, current understanding suggests an interaction between genetic and environmental factors as contributors [3,4].

Physical activity (PA) has a very important role and exerts a significant impact in patients with PD. Numerous evidence point to the remodelling and neuroplastic power of PA, including the role of muscle secretory activity in neurodegenerative diseases [5]. Further, PA may be responsible for both systemic [5,6] and neural [7,8] plasticity, defining the concept of *neural systemic dual plasticity* (NSDP) [9]. In the design of this protocol, we aim to determine the effect of exercise at the cellular and molecular level and having as its epicentre mitochondrial function related to changes in gene expression and functionality of mitochondria [10,11]. During PA, a number of myokines and metabolites are released into the bloodstream, many of which can cross the blood-brain barrier (BBB) and exert effects on the central nervous system, in this case the effects of exercise at the systemic level begin as secondary plasticity, that follow different signalling pathways and by passing the BBB has the potential to become primary plasticity [9].

Research has demonstrated that PA not only prevents cognitive decline and the risk of dementia in older adults [12] and other neurodegenerative diseases [13,14] but also mitigates motor deficits, increases new neuron formation, ameliorates neurological impairments, and helps counteract age-related neuronal loss [15]. Further, recent studies have indicated that intensive and cognitively demanding programs are capable of inducing plastic brain changes in individuals with PD [16]. PA plays a crucial role in modulating cortical activity among patients with PD [17]. It has been described that the activation of cortical areas during PA can be attributed to the increased intracerebral blood flow induced by exercise. A study evaluated the motor symptoms of PD using the Unified Parkinson’s Disease Rating Scale-III (UPDRS-III) and compared three groups of patients. Two groups participated in different PA programs, while the third group underwent physiotherapy. Remarkably, both PA groups exhibited a significant improvement of 27.5% in motor symptoms as assessed by UPDRS-III pre and post intervention, whereas the physiotherapy group showed a modest improvement of 2.9%. Notably, all the three groups demonstrated enhanced functional capacity at the end of the intervention [18]. These findings underscore the substantial benefits of PA in ameliorating motor symptoms and enhancing functional outcomes in individuals with PD. On the other hand, the relationship of PA with specific neurological benefits [19], highlights that PA activates a series of processes responsible for maintaining and protecting nerve cells, a phenomenon often referred to as physiological neuroprotection systems. PA favors the production of compensation mechanisms through a reorganization of damaged neural circuits [20].

In healthy subjects, the beneficial effects induced by PA are mediated by improved mitochondrial function and mitophagy [21]. This is relevant, since mitochondrial dysfunction is a key phenomenon associated with early PD, which occurs before the onset of motor symptoms [22]. PA has been shown to improve mitochondrial respiration, thereby affecting adenosine triphosphate (ATP) production and overall mitochondrial function. Indeed, both acute exercise and resistance exercise increase breathing and respiratory control index [23,24].

Moreover, skin fibroblasts from patients with PD have emerged as a valuable resource for studying this neurodegenerative disorder and identifying potential biomarkers [25]. Since acquiring human brain samples is only feasible *postmortem*, several researchers have turned to the easily accessible peripheral samples, such as skin fibroblasts. These studies emphasize that fibroblasts, originating from the same genetic lineage as neurons [26], can serve as reliable indicators of cumulative cellular damage related to the age and patient-specific habits [27]. The relevance of skin fibroblasts as a surrogate model in this research lies in their ability to mirror specific cellular deficiencies commonly observed in nigral neurons. These deficits serve as indicators the neurodegenerative process of PD and can be detected in fibroblasts from PD patients [27]. Hence the importance of fibroblasts as a surrogate model for physiopathological processes, as it would allow mitochondrial function to be correlated with clinical measures, such as motor function by means of the MDS-UPDRS III scale [28]. Our recent studies examining skin fibroblasts from PD patients have revealed metabolic and mitochondrial abnormalities [23], establishing a clear link between mitochondrial dysfunction in skin fibroblasts samples from PD patients and PD pathogenesis. These investigations demonstrated decreased oxygen consumption rate (OCR), proton leak, maximal respiration, respiratory replacement capacity, and OCR-linked ATP levels in skin fibroblasts from patients with PD, providing clear evidence of compromised mitochondrial function.

The aim of this clinical study is to explore the impact of PA on metabolic remodeling, utilizing a less invasive model to investigate PD and its pathogenic mechanisms. Specifically, the study aims to examine fibroblasts derived from PD patients, which serve as an accessible and informative model to gain insights into the underlying deficits affecting neurons. By investigating the beneficial mechanisms associated with PA and studying the pathophysiological alterations within fibroblasts, this research holds the potential to deepen our understanding of PD and facilitate the development of novel diagnostic tools, and revolutionary therapeutic interventions applicable to clinical practice.

## Methods and analysis

### Study design

PARKEX trial is a randomized clinical trial with an open design (registered at clinicaltrials.gov; NCT05963425). It is a non-pharmacologically interventional study, with a total of twenty-four patients. Screening and assessment visits are occurring in both neurology and PA laboratory settings.

A SPIRIT schedule and overview of the study design can be found in Figure 1 and Figure 2 below.

**Figure 1.**
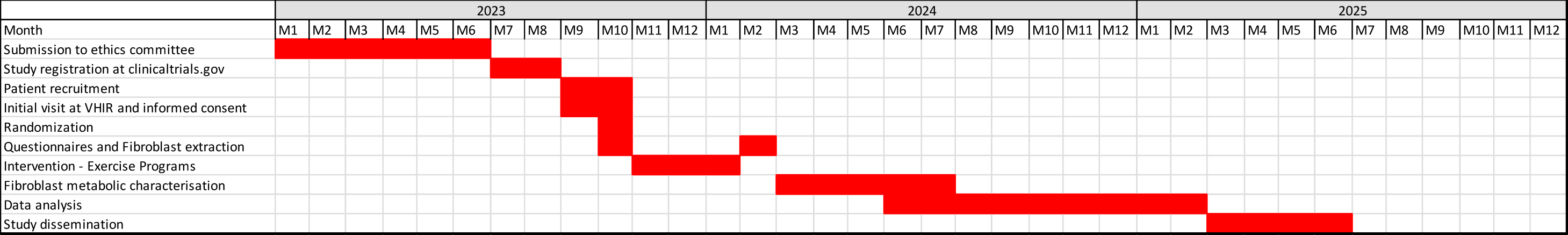
SPIRIT schedule for the PARKEX study.

**Figure 2.**
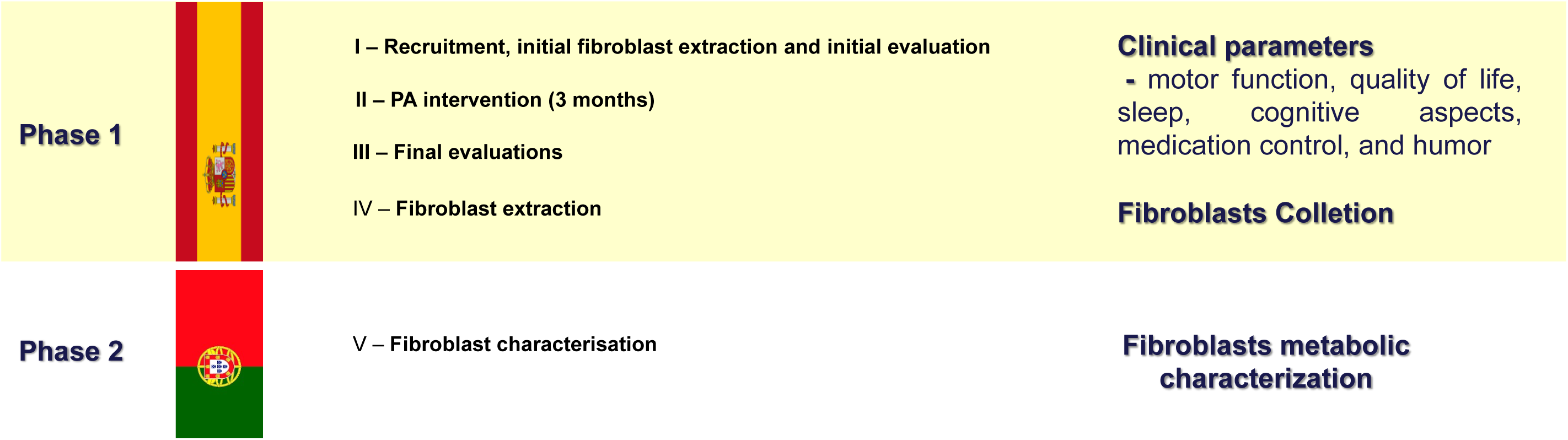
Comprehensive overview of the PARKEX study design.

This protocol has been developed following the Standard Protocol Items: Recommendations for Interventional Trials (SPIRIT) checklist [29].

### Subject Recruitment

Participants for the study will be recruited by the Grup de Malalties Neurodegeneratives of the Vall d’Hebron Institut de Recerca (VHIR), Barcelona, Spain. To ensure the minimum required sample size of 24 patients, as stipulated in the project, the VHIR researchers will leverage their connections with the Catalan Parkinson’s Association and other Movement Disorders Units in prominent hospitals across the state. Participants who meet the inclusion criteria and sign the informed consent will be recruited and randomized.

### Sample Size Calculation

The sample size for this study was determined using the GRANMO program with the model of observed proportions with respect to a reference. For the calculation it has been taken into account that the “maximal respiration” is the key indicator to determine the effects of PA on mitochondrial function, because the entire respiratory chain is forced to the maximum. The calculation has been made accepting an □ risk of 0.05 and a beta risk of less than 0.2 in a two-sided contrast. Based on preliminary data, there is a difference of “maximal respiration” equal to or greater than 0.263 units [23]. It is assumed that the proportion in the reference group is 0.019 in people older than 50 years [30]. Being an interventional study, the percentage of necessary replacements has been predicted to be 20%.

### Eligibility criteria

To be included in this study, subjects must meet specific criteria. These criteria include having a medical diagnosis of idiopathic PD, falling within stages of I-III on the H&Y scale in the ‘on’ phase, and demonstrating a good cognitive status with a score 26 or higher on the Montreal Cognitive Assessment (MoCA). Participants must be between 50 and 70 years old and capable of walking independently for at least six minutes. In addition, it is important that the subjects have not made any changes to their medication regimen in the last month. The International Physical Activity Questionnaires (IPAQ) will be used to normalize the groups based on the basal levels of PA [31]. Once the subjects are selected, they will receive a detailed explanation of the research procedures, and the subjects will voluntarily express their consent to participate in this study, signing the informed consent.

Certain criteria will lead to the exclusion of subjects from participation in this study. These criteria encompass the presence of any pathology other than idiopathic PD, cognitive impairment with a MoCA score below 26 points, uncontrolled cardiovascular disease, diabetes, visual impairment, or recent musculoskeletal disorders in the upper or lower extremities that may interfere with balance and locomotion. Additionally, individuals currently undergoing another therapeutic exercise protocol or who have undergone surgery aimed at influencing a specific PD symptom will not be eligible to participate.

### Randomization

All patients who provide their consent to participate in the study and fulfil the inclusion criteria will be randomized. A sample block randomization method will be employed, dividing the participants in three groups: two intervention groups and one control group. The allocation of individuals to intervention or control groups will be blind to assessors and participants. An independent investigator, uninvolved in the study, will perform the random assignment of subjects into the three groups. This will be done using a computer-generated random allocation table, stratified based on the PD stages (i.e., according to H&Y classification stages I, II, III), basal physical activity levels, sex, and age.

### Intervention

Two different programs are designed in order to investigate the effect of PA on mitochondrial function in skin fibroblasts of patients with PD.

The first group (n=8) will undergo basic physical training (BPT) focused on strength and resistance. The second group will engage in BPT combined with functional exercises (BPTFE), which includes cognitive and dual task training. A third group, the sedentary group, will serve as the control. The intervention in the two PA programs will last 3 months.

The interventions will be conducted in group settings, with 8 patients per group, and each session will last for 60 minutes. The frequency of the sessions will be 3 times a week. The intervention programs will be structured into 3 meso-cycles of 4 weeks. During the initial two weeks, the workloads will be maintained to facilitate the adaptation period. For the BPTFE program, in addition to adapting to the load, participants will also need to adapt to complex motor execution tasks. Starting from the third week, the workload will progressively increase.

In both PA programs, exercises will target the large muscle groups, with a component/emphasis on the eccentric phase of concentration. The *kBox4* Platform, a device designed to maximize performance and training outcomes, will be used to enhance the eccentric phase of the exercises. This device, provided by Exxentric AB (Stockholm, Sweden), a company specialized in innovative and evidence-based training equipment and methods, will be loaned for the study. *kBox4* is controlled by a specific program, *Kmeter*, which records and stores all the information (duration, intensity, repetitions, etc.) of each movement. Exxentric *kBox4* will be adapted with special harnesses, insurances on the wall and in front of support bars, according to the needs of the patients. In addition to the major muscle groups, specific attention will be given to the key muscles involved in the gait cycle, such as the tibialis anterior, medial gastrocnemius, rectus femoris, and hamstrings, from a biomechanical perspective. This aspect will be common to both programs, with the only difference being that the BPT program will have a greater workload due to the division of activity time between functional exercises and Dual Task Training in the BPTFE program.

### Experimental groups

#### Intervention group 1 – Basic Physical Training (BPT)

This PA program will be based only on the work of BPT, in which specifically and mainly the Strength (S) and Resistance (R) will be worked, but also flexibility.

#### Intervention group 2 – Basic Physical Training with Functional Exercises (BPTFE)

In addition to the BPT program (which includes the transverse focus on strength and resistance exercises), this intervention group will incorporate functional exercises that involve dual task training. The dual task training can encompass both motor-motor and motor-cognitive activities. This means that coordinated exercises will be performed, with cognitive activities introduced at the extremes of the movement. This approach ensures engagement of both physical and cognitive abilities during the PA sessions.

#### Control group – Sedentary (Sed)

Participants in the control group will maintain their regular daily routines throughout the study period. Weekly interviews will be conducted by the researchers to ensure that their routines remain unchanged and to minimize any effect due to the interaction with the researchers. After the final evaluation, at the end of the study (6 months), this group will receive 3 months of PA based on the program that has obtained the most favorable outcomes, in terms of symptoms management and quality of life.

### Outcome assessment and measurement procedures

The primary outcome of the PARKEX study is the mitochondrial (dys)function, while secondary outcomes include motor function, quality of life, sleep, cognitive aspects and mood. Additionally, the study aims to perform the metabolic characterization of cutaneous fibroblasts from PD patients, evaluating the effects of PA-induced metabolic remodeling on oxidative stress, mitochondrial quality control, mitochondrial DNA copy number, mitochondrial protein expression and transcript levels in skin fibroblasts of PD patients.

To gather pre-intervention and post-intervention clinical parameters, the following experimental design procedures will be followed in 2 different phases (see Table 1, Table 2 and Figure 3):

**Figure 3.**
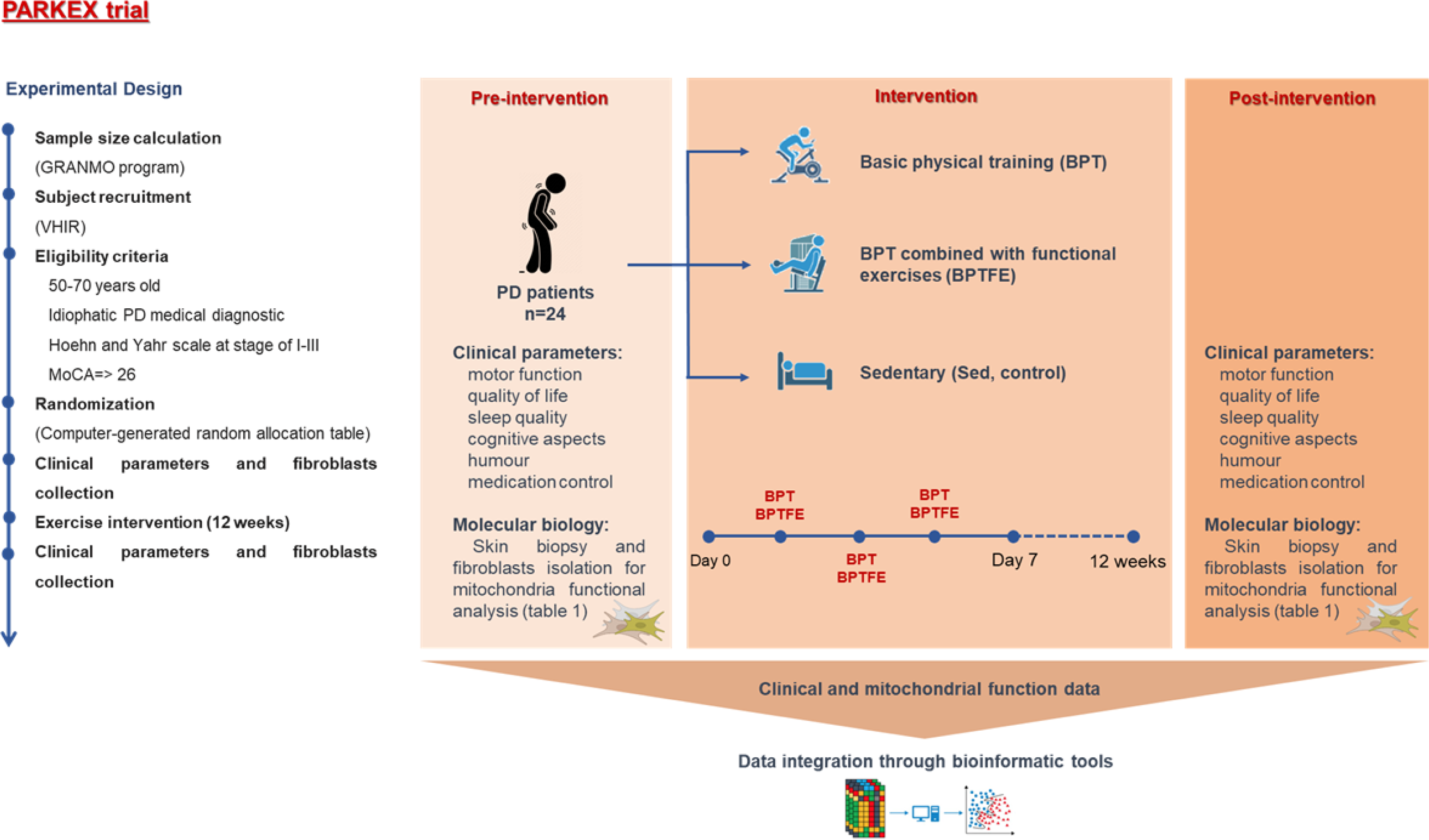
Graphical representation of PARKEX study design, highlighting the interventions and respective experimental groups, timing of sample collection, and analyzed endpoints.

**Table 1:** Phase 1 in Spain: Secondary outcome measures and their assessment instruments.

| Secondary outcome measures | Instruments to assess the study variable |
| --- | --- |
| **Functional Assessment - Balance** | TINETTI scale and BERG Balance Scale |
| **Functional Assessment** | Six-minute walk test (6MWT) |
|  | 1-minute sit-to-stand test (STS). Times you get up and sit down, in a chair |
|  | Timed Up and Go (TUG). Time to get up from a chair, walk 3 m, turn around, walk backwards and sit down |
| **Motor function** | Movement Disorder Society - Unified Parkinson's Disease Rating Scale (MDS-UPDRS I-III) |
| **Cognitive aspects** | Montreal Cognitive Assessment (MoCA) |
| **Quality of life** | Parkinson's Disease Questionnaire (PDQ-39) |
| **Humour (Profile of Mood States)** | BDI-2 Beck's Depression Inventory |
| **Sleep quality** | Parkinson's Disease Sleep Scale-2 (PDSS-2) |
| **Non-motor symptoms** | MDS-NMS scale, Scopa-Aut |

**Table 2:** Phase 2 in Portugal: Assessment of surrogate patterns for mitochondrial (dys)function in patients’ fibroblasts using molecular biology techniques.

| Primary outcome measures | Specific objectives | Description | Mitochondrial function analysis |
| --- | --- | --- | --- |
| Evaluate the effect of PA on mitochondrial remodeling in skin fibroblasts from patients with PD, namely in the mitochondrial maximal respiration | Assess the effects of metabolic remodeling on mitochondrial oxygen consumption rate, oxidative stress, mitochondrial quality control, mitochondrial DNA copy number, mitochondrial proteins and transcripts | Characterize the effects of PA remodeling on fibroblast from patients with PD, in terms of mitochondrial biogenesis, oxidative stress, and mitochondrial quality control | - Determination of oxygen consumption rate (OCR) and extracellular acidification rate (ECAR) using a Seahorse XFe96 Extracellular Flux Analyzer (Agilent Technologies, Germany).<br>- qRT-PCR (using SsoFast Eva Green Supermix, in a CFX96 real time-PCR system (Bio-Rad, Hercules, CA, USA)) for transcripts of interest related to mitochondrial biogenesis, dynamics, epigenetic regulation, oxidative phosphorylation, oxidative stress (SOD1, SOD2, NFE2L2), quality control mechanisms, including autophagy and ubiquitin-proteasome system (UPS).<br>- Regulation and abundance of mitochondrial proteins by immunoblotting<br>- Enzymatic activities (ex, mitochondrial complex I and citrate synthase) |

### Baseline assessment

Each participant will need to sign an informed consent prior to being interviewed. The interview will include a comprehensive medical history review and a complete anamnesis including the collection of medication control, gathering data on their current health status and also the chronology of their specific pathology. The clinical history and neurological examination will be prepared to confirm inclusion-exclusion criteria, assessing motor strength, muscular tone and pathological reflexes. Familiarization sessions will also be conducted before the intervention. In this session, the participants will be provided with a smart band (Amazfit Band®) in order to normalize the basal levels of PA that the patients may have throughout the study.

Both intervention and control groups will undergo a baseline evaluation, including functional assessment, PA tests, motor assessment, medication control, health assessment and skin biopsy (small tissue samples) for fibroblast extraction from the skin of the neck.

#### 1. Baseline evaluation

A battery of tests will be performed (Figure 2), encompassing a complete neurological examination to confirm inclusion-exclusion criteria using the MDS-UPDRS questionnaire; psychological tests aimed at assessing cognitive aspects include the MoCA questionnaire; quality of life (using the PDQ39 questionnaire), mood (using the BDI-2 questionnaire), balance (TINETTI), sleep (PDSS test), and dysautonomia (SCOPA-AUT test). The 6-minute walk test (6MWT) to evaluate the functional capacity and endurance of participants will be conducted according to standardized guidelines. These tests and questionnaires will be evaluated pre– and post-intervention (at 3 months) and 3 months after the end of the intervention (6 months after the initial evaluation).

#### 2. Health assessment

A comprehensive health assessment will be conducted by the Neurodegenerative Diseases Research Group at VHIR to compile a complete medical history. Health parameters (heart rate, blood pressure, anthropometric measurements, height, weight, BMI) will be determined before and after intervention.

#### 3. Skin fibroblast extraction

Specialized neurologists from the Neurodegenerative Diseases Research Group at VHIR will perform the biopsy for skin fibroblast isolation. This will allow us to investigate patients’ fibroblast’s mitochondrial function, which was being described as impacted in neurodegenerative diseases such as PD, and thus enabling the opportunity to verify the effect of applying a PA program in these patients.

Skin fibroblast biopsies will be collected before and after the intervention in all three groups. The collected cells will then be cultured, expanded, and frozen at VHIR. Subsequently, the cells will be sent to the Center for Neuroscience and Cell Biology at the University of Coimbra, Portugal for further functional and molecular analysis.

### Mitochondrial analysis

The analysis of mitochondrial function and metabolism in skin fibroblasts from patients will encompass various parameters. Metabolic fluxes of the cells will be assessed using the Seahorse XF^e^96 Extracellular Flux Analyzer [23]. Additionally, mitochondrial membrane potential and morphology will be evaluated using TMRM staining, while cellular oxidative stress will be measured using hydroethidine (HEt) and CM-H2DCFDA. Moreover, ATP levels will be determined using a high-throughput luminescent method [37]. The expression of relevant transcripts associated with mitochondrial biogenesis, stress responses, oxidative phosphorylation, and auto(mito)phagy will also be evaluated [23]. This includes enzymes such as superoxide dismutase, catalase [38], glutathione reductase, parkin and PINK1 proteins, the mitochondrial transcription factor A (TFAM), the regulator of mitochondrial biogenesis (such as PGC1-α), and metabolism (such as AMPK-α) [37]. The evaluation of these parameters aims to unravel the molecular mechanism through which PA promotes mitochondrial metabolism remodeling in PD patients. Furthermore, this analysis seeks to identify new intervention targets for PD and demonstrate, for the first time, that the metabolic and mitochondrial beneficial effects of PA in PD patients can be detected through observable biochemical changes in their skin fibroblasts. By uncovering these biochemically-detectable alterations, we can gain valuable insights into the specific mechanisms underlying the metabolic and mitochondrial improvements resulting from PA, providing a foundation for targeted interventions in PD.

### Statistical analysis

Data analysis will be performed using SPSS v.20 (IBM SPSS Statistics). Regarding descriptive analysis, all the variables of the sample will be analyzed by means of relative and absolute frequencies, as well as measures of dispersion and central tendency frequencies. Repeated measures analysis will be performed using the appropriate mixed effects model, accounting for homogeneous or inhomogeneous samples. This analysis will determine the possible changes between the different moments of the intervention and to evaluate which types of intervention yield the greatest physical benefits, whether in terms of mitochondrial function, physiological factors, quality of life, or cognitive and emotional aspects. Spearman’s rho and Pearson’s r will be used to evaluate the association between two variables that have ordinal categories. The normality of the distribution of the results for each group’s results will be assessed using the Shapiro-Wilk normality test, with a significant threshold set at α=0.05. If the data follow a normal distribution, a parametric paired t-test will be conducted. Otherwise, the Mann-Whitney test will be used. Statistical significant differences will be considered for statistical test values with p<0.05.

## Ethics

The research project has received approval from the Research Ethics Committee of the Faculty of Psychology and Education and Sports Sciences (Blanquerna, Universitat Ramon Llull) on 27/01/23 (2021008D), as well as from the Ethics Committee for Research with Medicines at Vall d’Hebron University Hospital (PR(AG)574/2021). Participants will sign informed consent prior to the participation of the trial.

## Discussion

Accumulating evidence supports the potential benefits of PA for patients with PD, including both general health improvements and disease-specific effects. However, the exact mechanism connecting skeletal muscle-increased activity and mitochondrial remodeling in PD are poorly elucidated. The PARKEX study is the first clinical trial that aims to evaluate the effects of two PA programs on skin fibroblasts mitochondrial function from patients with PD, as well as their impact on motor function, quality of life, sleep, cognitive and mood aspects. The 12-week implementation of the BPT and BPTFE programs is expected to positively impact relevant clinical aspects of PD by enhancing systemic mitochondrial function, restoring mitochondrial metabolism, and influencing gene expression patterns. These improvements are anticipated to translate into potential neuroprotective effects. Additionally, the PARKEX clinical trial will provide insights into the degree of mitochondrial function improvements in PD through a comparative analysis of BPT and BPTFE programs and aims to unveil, for the first time, the biochemically-detectable changes in skin fibroblasts from PD patients that reflect the metabolic and mitochondrial benefits of PA. This crucial observation will demonstrate the tangible and measurable effects of PA on mitochondrial function at the cellular level. Understanding the regulation of the mitochondrial function by PA in PD holds significant promise in defining interventions to delay disease onset and developing new therapeutic approaches not only for PD but also for other neurodegenerative disorders.

## Trial status

As of the time of this publication, the study is actively underway in accordance with protocol version 1.3, dated May 17, 2023. Recruitment of participants started in September 15^th^ and is anticipated to be completed by October 2023. The intervention phase and the last follow-up assessments are expected to be concluded by January 2024. The study is progressing according to the planned timeline, and data collection and analysis will commence thereafter to generate meaningful results.

## Supporting information

S1 Checklist. SPIRIT 2013 checklist: Recommended items to address in a clinical trial protocol and related documents S2 Clinical trial protocol, v4, V4; December 05 2022

## Data Availability

No datasets were generated or analysed during the current study. All relevant data from this study will be made available upon study completion

## Acknowledgments

The authors would like to thank all PD patients who will contribute to the performance of the study and the Associació Catalana per al Parkinson for their invaluable help in diffusion of the study and the subject recruitment.

## Author contributions

J.C.M., C.M.D., L.B., and J.M. wrote the original draft preparation of the manuscript. M.A., E.G.R., S.E.C., A.L., J.H.V., M.M.V., M.G.-G., J.M. and S.P.P. wrote, reviewed and edited the manuscript. All authors critically revised the final form of the manuscript. All authors have read and agreed to the published version of the manuscript.

